# Testing the feasibility of a national respiratory swabbing tool in FluSurvey to support pandemic preparedness in England, 2025-26

**DOI:** 10.64898/2026.09.09.26362615

**Authors:** Hannah S Wolmuth-Gordon, Meera Lachhani, Richard Pebody, Gayatri Amirthalingam, Rebecca E Green, Gavin Dabrera

## Abstract

**Background:** FluSurvey is a participatory surveillance system, through which participants report weekly respiratory symptoms, healthcare use and social contacts. To support rapid public health assessment we developed a respiratory self-swabbing feature. We aimed to assess the feasibility of self-swabbing among symptomatic FluSurvey participants.

**Methods:** FluSurvey self-swabbing was deployed in England between January-March 2026. Adults reporting recent Acute Respiratory Infection symptoms were eligible for a self-swabbing Covid-19, RSV and Influenza PCR test which was returned to the laboratory by post. Participants were sent optional feedback surveys. We described swabbing interest, timeliness and participant demographics. We evaluated participant feedback to improve the process.

**Results:** There were 234 eligible illnesses during the study period; 45.7% (107/234) requested test kits and 93.5%(100/107) test kits were returned. Of these, 97.0%(97/100) had valid results; median participant age was 61.74 years (IQR:47.4-67.4), 68.4% were female and 90.0% were of white ethnicity. All nine regions in England were represented. Timeliness was high; the median time from symptom onset to specimen date was 4.0 days (IQR:3.0-5.0). Of participants completing feedback surveys, 96.7%(59/61) would consider participating in self-swabbing again.

**Conclusions:** We developed a tool to test individuals, often not presenting to healthcare, for respiratory viruses using an established community-based surveillance system. This tool can be rapidly deployed and adapted to public health need, to assess community-based emerging respiratory or pandemic infections. The high return rate and positive feedback suggest continued engagement of the cohort in respiratory virus swabbing.

## Introduction

Participatory surveillance systems can offer near-real time monitoring of community respiratory illness, including of those not attending healthcare. This can complement insights from traditional surveillance systems and has the benefit of being cost-effective, flexible and scalable (World Health Organization, 2024). Due to the two way interaction between public health bodies and participants, these systems can also be used to communicate public health information, which can be tailored to specific participant groups (Smolinski et al., 2017).

FluSurvey was established in 2009 and is an internet-based UK participatory surveillance system (FluSurvey, 2026) that monitors self-reported respiratory illness symptoms in the general population. In addition, participants report weekly social contact behaviour, absenteeism and healthcare-use. Combined with other surveillance systems, FluSurvey enables the monitoring of ‘influenza like illness’ (ILI) and other respiratory symptoms in the community (UK Health Security Agency, 2026b). However, the lack of specificity on the causative pathogen limits our capacity to understand circulating respiratory viruses in the community and detect emerging respiratory viruses (Mellor et al., 2025).

Self-swabbing has previously been available for FluSurvey participants for limited periods and was last available in 2020 (Edelstein et al., 2021; Wenham et al., 2018). Other similar internet-based participatory surveillance systems have recently deployed self-swabbing, such as the Netherland’s *Infectieradar* and Australia’s *FluTracking* surveillance systems (Esneau et al., 2026; Smit, Carstens, Han, Bulsink, Bakker, et al., 2025). Since self-swabbing was last available for FluSurvey participants, the FluSurvey web platform has significantly developed to enable greater automation, flexibility and scalability. Leveraging these digital developments, we additionally aimed to assess whether the current FluSurvey cohort were interested in self-swabbing and whether nose and throat swabs were acceptable amongst participants.

To support rapid public health assessment and monitoring of emerging respiratory viruses, we developed a FluSurvey respiratory self-swabbing tool for adult participants reporting acute respiratory infection symptoms. In winter 2025-26, the self-swabbing feature was trialled in England to test its feasibility to monitor community respiratory virus infections. We aimed to assess participants’ interest in self-swabbing, test kit response rates and the timeliness of test results.

## Methods

### Participants and study design

FluSurvey is a web-based participatory surveillance system in the UK that was established in 2009. FluSurvey collects data on participant demographics and lifestyles, in addition to weekly symptoms, healthcare seeking behaviour and social contact patterns during the autumn and winter. Participants are over 18 and live in the UK. In addition to themselves, participants may also report on behalf of other household members, including children. When participants register for FluSurvey, and at the start of every season, they complete a ‘profile’ survey that includes questions on demographics, lifestyle and risk factors of respiratory virus illness. Each week participants complete a ‘weekly’ survey that collects information on symptoms, healthcare use and social contact behaviour. Please see Green et al. (2026) for a detailed methodology of the FluSurvey study design and the supplementary material for the profile and weekly survey questions.

In winter 2026 a FluSurvey respiratory self-swabbing feature was piloted. Self-swabbing was available to FluSurvey participants that met the ARI case-definition (EU Commission, 2018) and the following criteria when they completed their weekly survey:

- Lives in England
- Aged at least 18 years
- ARI case definition: A sudden onset of at least one of the following symptoms: sneezing/cough/runny or blocked nose/sore throat/shortness of breath
- Symptom onset date within 3 days
- No prior swab code assigned in the previous 7 days

Participants who were eligible to participate in the self-swabbing pilot were asked whether they were interested in participating, and if they were, whether they consented to participate in the pilot. Participants who consented to participate were assigned a ‘swab code’ that they automatically received by email. To receive a free postal self-swab testing kit, participants were requested to redeem a swab code at Saving Lives, Take a Test (https://takeatestuk.com/). When participants redeemed their swab code they completed a Saving Lives questionnaire, in which name, sex, date of birth, NHS number, date of symptom onset, address, and contact details are provided. Participants received a nose and throat swab kit with instructions and returned this to the laboratory by post. The sample was tested in a laboratory for RSV, SARS-CoV-2, influenza A and influenza B by multiplex PCR. Positive influenza A samples were further subtyped into A(H1N1pdm09) and A(H3N2). The Saving Lives questionnaire and test results were securely transferred to the UKHSA FluSurvey team. Participants received test results by text message within 2 weeks. Approximately three weeks after participation, participants were sent a feedback survey to aid study evaluation. Feedback survey questions are in the supplementary material.

### Data processing

Data processing and analysis were conducted in R and R Studio version “Apple blossom” (Posit team, 2026; The R Foundation for Statistical Computing, 2024).

### FluSurvey dataset

FluSurvey weekly survey data was filtered for when self-swabbing was active (see Table S1). FluSurvey profile and weekly data are pseudonymised, with participants assigned a participant ID. Profile and weekly survey data were joined using participant ID. Participants update their profile survey throughout the season to reflect changes such as vaccination status. The linked FluSurvey dataset was deduplicated based on which profile survey was completed closest in advance of participants’ weekly survey.

Age was calculated as the time between date of birth and the date weekly survey was submitted. Sample sizes of many of the ONS ethnic groups included in the FluSurvey profile survey were small and therefore, ethnicity was grouped into larger aggregations (white, other and unknown when missing).

### Test result dataset

The FluSurvey survey dataset was joined to Saving Lives survey and test results using swab code. This dataset was linked to additional laboratory data by NHS number and when this was not available forename and surname. The dataset was enriched by linking to the ONS postcode directory (Office for National Statistics, 2026) by postcode, which was provided by participants in their Saving Lives questionnaire. This enabled geographical distribution of participants to be described.

### Data Analysis

We used R and RStudio to calculate descriptive statistics relating to participant demographics, test results, eligibility and timeliness of test results. Plots were created using ggplot2 (Wickham, 2016). To evaluate the pilot results in the context of additional respiratory surveillance data we examined influenza, RSV and Covid-19 case positivity in the Respiratory DataMart sentinel laboratory surveillance system (UK Health Security Agency, 2025).

We described demographics of the eligible FluSurvey cohort using FluSurvey data and participants who received test results using Saving Lives survey data. We examined healthcare use amongst eligible participants and those that received test results. We calculated several variables to estimate timeliness: the number of days between symptom onset and requesting a swab in FluSurvey, days between requesting a swab in FluSurvey and laboratory specimen date, and days between requesting a swab in FluSurvey and laboratory received date.

### Privacy and ethics

Ethical approval from the UKHSA ethics board was obtained for the study (ref: NR0433). Participants read and accept a privacy notice when they sign up for FluSurvey (https://flusurvey.net/legal/privacy). Self-swabbing participants were shown an additional privacy notice and consented before participating in the study (https://flusurvey.net/legal/privacy-postal-swabbing).

## Results

### Overall

FluSurvey self-swabbing was available to participants for nine weeks from 19/01/26 – 20/03/26. To enable the close monitoring of the new self-swabbing feature during this trial period, swabbing was available at specific times (Table S1).

234 reported illnesses were eligible for self-swabbing, 107 requested a swab kit, and 100 test results were received, of which 97 were valid. Fig. 1 shows the number of participants at each stage of the process.

**Table 1.**
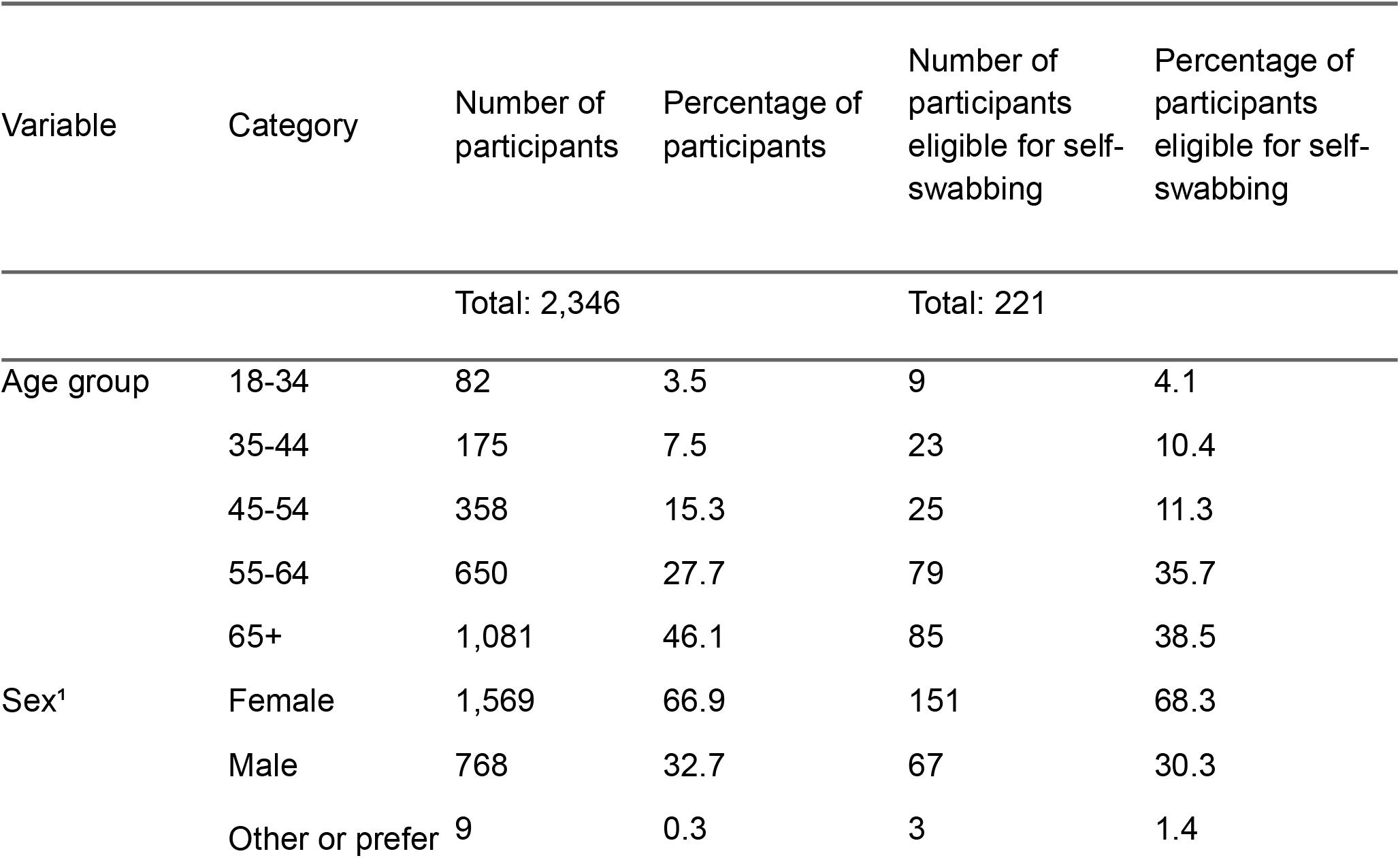
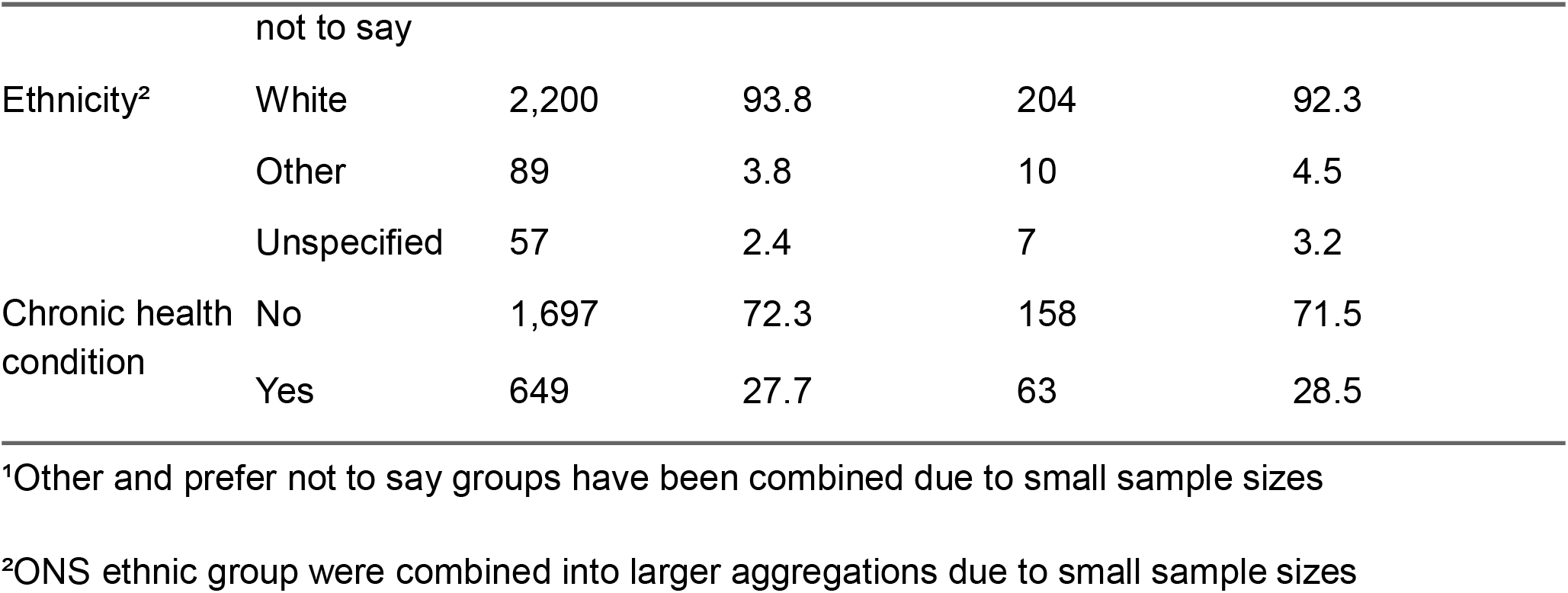
Characteristics of FluSurvey participants that live in England and are older than 18 years and of those who were eligible for self-swabbing. Only participants that contributed to FluSurvey when self-swabbing was available are included.

**Fig 1.**
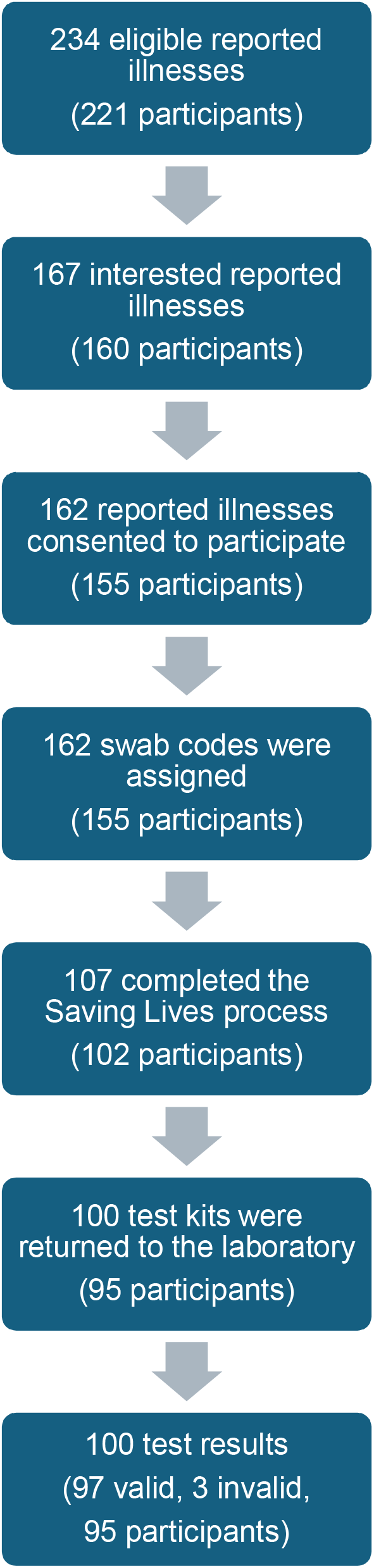
Flow diagram showing the number of reported illnesses and participants at each stage of the FluSurvey self-swabbing process.

### Eligibility and interest in swabbing

During the entire study period, 234 reported illnesses (relating to 221 participants) met all criteria and were eligible for swabbing. Participants reporting eligible illnesses were presented with additional information on the self-swabbing study. Of these eligible illnesses, 71.4% (167/234 illnesses, 160 participants) were interested in participating and 28.6% (67/234 illnesses, 61 participants) were not interested. Of the 167 eligible and interested illness reports, 97% (162/167 illnesses, 155/160 participants) consented to participate in the study and 3% of illnesses did not consent (5/167 illnesses, 5/160 participants; Fig. 1).

Of the 221 eligible participants, 83.2% (184) had previously expressed an interest in swabbing in their profile survey, 13.1% (29) had expressed potential interest and 3.6% (8) were not interested.

### Swab codes & test kits

162 swab codes were assigned to 155 participants. Five participants requested multiple swab codes. The highest number of swab codes requested was during the week commencing 16^th^ March, the final week of the trial period.

107 swab codes were registered with Saving Lives, with participants completing the process to receive a testing kit in the post. 93.5% (100/107) test kits were returned to the laboratory for testing (Fig. 1).

### Demographics & geography of participants with test results

The median age of participants from which we received test results was 61.74 years (IQR: 47.4 – 67.4 years) and 68.4% (65/95) were female and 31.6% (30/95) were male. The majority of participants were of white ethnicity (89.5%, 85/95) and 7.4% (7/95) participants were of another ethnicity. Participants were from all 9 regions in England (Fig. 2). London had the highest number of participants, followed by the South East of England. Combined, these represented 41.1% (39/95) of participants.

**Fig 2.**
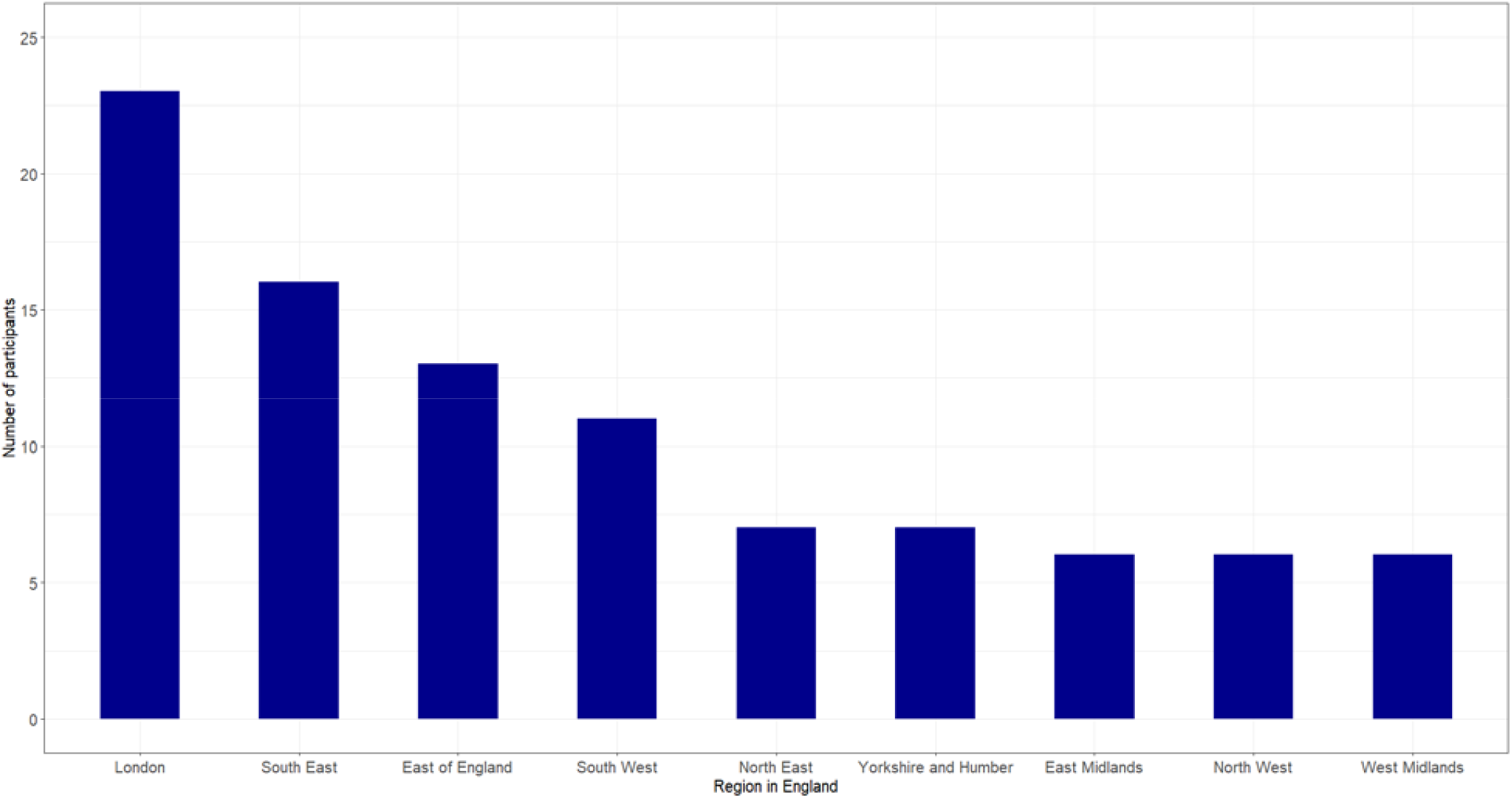
Number of participants that received test results and the region in England of residence.

### Healthcare use

Reported healthcare use was low across both eligible illness reports (1.3%, 3/234) and for illness reports that requested a swab code (1.2%, 2/162).

### Timeliness of swab codes and test results

The median number of days between symptom onset and firstly, requesting a swab test was 2.4 days (IQR: 1.5-2.6) and secondly, the specimen collection date was 4 days (IQR: 3-5). The median number of days between requesting a swab test and the specimen date was 1.5 days (IQR: 1.2-2.5). The median number of days between requesting a swab test and the laboratory receiving the sample was 5.4 days (IQR: 3.8-6.4). Please note that fewer samples were processed by the laboratory during the weekend (Fig. S1).

### Test results

We received 100 test results, of which 97 were valid and 3 were invalid. Valid results included positive and negative Covid-19, Influenza and RSV results. UKHSA surveillance data demonstrate that the study period was at the tail end of peak influenza and RSV activity and during a period of low but stable Covid-19 activity (see Fig. 3).

**Fig 3.**
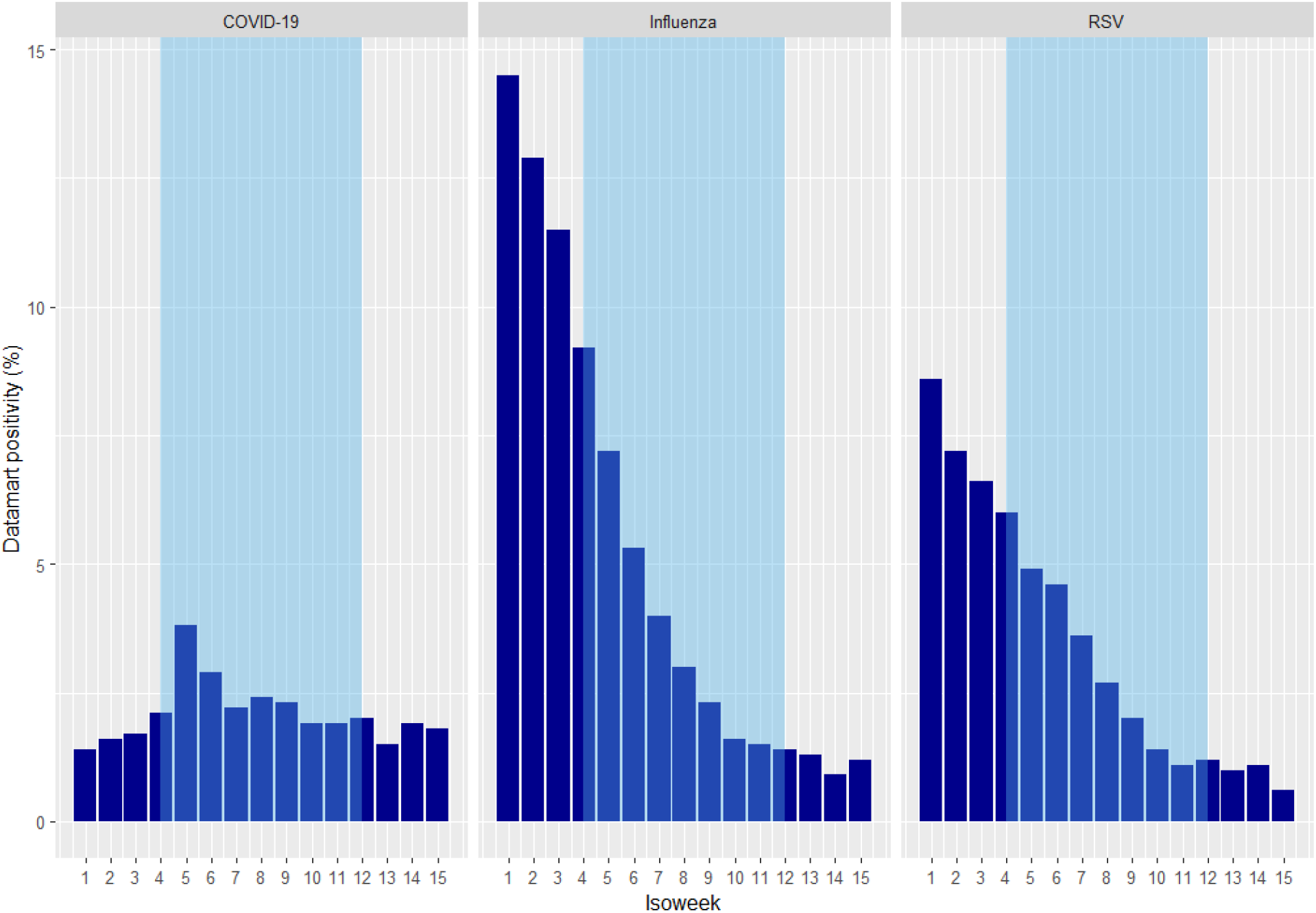
Datamart test-positivity for Covid-19, Influenza and RSV by isoweek. Datamart data is from the UKHSA Flu and Covid National Surveillance report published on April 16^th^ (UK Health Security Agency, 2026a). Shaded region shows the weeks that FluSurvey self-swabbing was available to FluSurvey participants. Please note, during these weeks swabbing was not available every day.

### Participant feedback

Of the 102 participants that requested a testing kit from Saving Lives, 72 participants completed feedback surveys. Of these participants 94.4% (68/72) strongly agreed or agreed that the Savings Lives process was easy to follow. Of these 72 participants, 61 received test results. 95.1% (58/61) strongly agreed or agreed that they understood their test results. 96.7% (59/61) strongly agreed or agreed that they would participate in self-swabbing for public health surveillance in the future.

## Discussion

We have demonstrated that respiratory self-swabbing in the community is feasible among symptomatic FluSurvey participants, including among cases not presenting to healthcare settings. Response rates were high, with 93.5% (100/107) of swab kits returned to the laboratory. The ease of requesting a kit (reported by 94.4% of participants) likely contributed to high engagement, in addition to the motivation of receiving their test results. It is also likely that interest and motivation are particularly high in this group because the cohort voluntarily participate and therefore, may be more engaged in the concept of self-swabbing for public health surveillance. The low number of participants attending healthcare during their eligible illness, emphasises that this surveillance system captures individuals outside of traditional healthcare based surveillance systems. In addition, feedback surveys indicate continued future interest and engagement in self-swabbing as has been found in other trials of participatory surveillance self-swabbing (Haussig et al., 2019; Wenham et al., 2018).

The timeliness was high, particularly the period between symptom onset and specimen date, for which the timeliness is important to ensure the virus is detectable in the sample (Ip et al., 2012). Estimates of timeliness are not comparable with that of the previous FluSurvey pilot, as participants received testing kits prior to symptom onset (Wenham et al., 2018). We obtained valid results from 97% of samples and detected Influenza, SARS-CoV-2 and RSV, indicating that samples were of sufficient quality for virus detection, highlighting the important of timeliness. Self-swabbing was deployed towards the end of the influenza season, during a period of low respiratory virus activity (UK Health Security Agency, 2026b). Therefore, we did not receive a large sample of positive test results and were unable to compare results with that of other surveillance systems to assess external validity. This pilot was not intended to yield a representative sample of the respiratory viruses in circulation at the time. If the pilot was repeated in future, deploying self-swabbing during a period of high respiratory virus activity would be more informative in the assessment of the accuracy of respiratory virus detection and prevalence estimates in the community. Furthermore, the sufficient quality of self-swabbing samples presents a valuable opportunity to collect virus samples for sequencing. Sequencing viral samples would increase our understanding of the variants in circulation and enable prompt identification of new mutations and variants.

Approximately 70% of the eligible cohort participated in self-swabbing, with the largest participant dropout rate occurring between eligibility and interest (28.6%). The high proportion participating in the pilot is encouraging, but approximately 10% lower than estimates of self-swabbing interest reported on registration. It is possible that in a public health emergency participants would be more interested in swabbing and this dropout rate may fall (Boettiger et al., 2026). Interest and participation in self-swabbing may also be higher during peak winter respiratory virus activity. Following expressions of interest, participant response rates remained high and kit return rates were higher than the 2014-15 FluSurvey self-swabbing pilot, at 93.5% (100/107) compared to 77% (41/66) (Wenham et al., 2018).

The sample of participants taking self-swabs were not representative of the adult population in England. Like many participatory surveillance systems, our sample was biased towards older, female participants of white ethnicity (Boettiger et al., 2026; Makhasi et al., 2026; Smit, Carstens, Han, Bulsink, de Bakker, et al., 2025). Targeting recruitment at underrepresented groups could help to mitigate this bias, however this is unlikely to achieve a fully representative sample. Developing quota sampling when allocating swab tests could make the sample of self-swabbing users more representative of the population, particularly when the number of test kits is limited. Further work would be needed to assess the feasibility of adults swabbing children in a future public health emergency, as we did not include participants under 18 years. We also did not test the feasibility of testing asymptomatic individuals, which may be necessary if greater awareness of population prevalence was needed.

In conclusion, we have demonstrated that FluSurvey self-swabbing is feasible, with high test kit return rates, adequate samples for virus detection and high levels of interest in future self-swabbing. This tool can be rapidly deployed and adapted according to public health need, for example to test for different respiratory viruses or target specific risk groups. This makes it a valuable surveillance tool for detecting emerging respiratory pathogens or in a pandemic, as well as flexibility to monitor seasonal respiratory activity in the future to inform situational awareness of community respiratory illness.

## Data Availability

Authors cannot make the underlying dataset publicly available for ethical and legal reasons.

