## Supplementary material for "Testing the feasibility of a national respiratory swabbing tool in FluSurvey to support pandemic preparedness in England, 2025-26"

**Table S1** Time self-swabbing was available to eligible FluSurvey participants (over 18 years and live in England). In week 4 the criteria for timing of symptom onset was changed from 2 to 3 days (shown by *).

| **Week of**  **swabbing pilot** | **Time swabbing available** | **Number of hours swabbing available** |
| --- | --- | --- |
| 1 | 19/01/26 10:00 – 19/01/26 15:00  20/01/26 12:00 – 22/01/26 16:00 | 57 |
| 2 | 26/01/26 00:00 – 29/01/26 12:00 | 84 |
| 3 | 02/02/26 00:00 – 05/02/26 12:00 | 84 |
| 4* | 08/02/26 00:00 – 10/02/26 12:10  12/02/26 11:09 - 12/02/26 15:45 | 64.5 |
| 5 | 16/02/26 9:00 – 16/02/26 16:00  17/02/26 9:00 – 17/02/26 16:00  18/02/26 9:06 – 19/02/26 16:00 | 45 |
| 6^+^ | 23/02/26 00:00 – 26/02/26 16:00 | 88 |
| 7 | 02/03/26 00:00 – 05/03/26 16:00 | 88 |
| 8 | 09/03/26 00:00 – 12/03/26 16:00 | 88 |
| 9 | 16/03/26 00:00 – 20/03/26 16:00 | 112 |

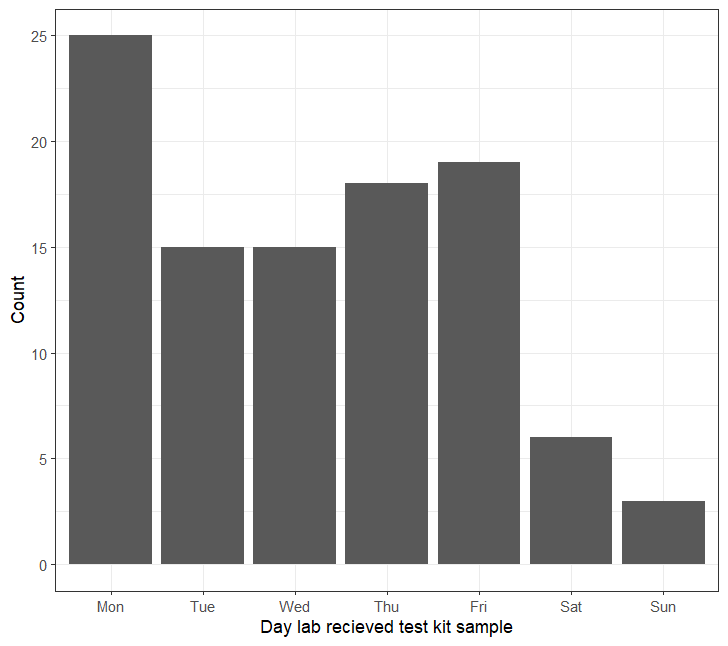

**Fig. S1** The number of test kits received by the laboratories on each day of the week.

### FluSurvey profile questionnaire

* indicates mandatory questions

1. For whom are you filling in this survey?*
   *If you are filling in the survey on behalf of someone else, then make sure that you have the consent of that person to do so.*

- Myself
- A member of my household
- Someone else

2. What is your sex?*

- Male
- Female
- Other (this will be updated to “Prefer not to say”)

3. What is your date of birth (year and month)?

- Year: 1920-2025
- Month: January-December

4. What is your current Country of residence?*

- England
- Scotland
- Wales
- Northern Ireland
- Isle of Man
- Channel Islands

5. What is the first part of your home postcode (the part before the space)?*

- Postcode:<first part of your postcode>
- I don't know/can't remember

6. What is your ethnic group?
   *This question is optional.*

- Asian, Asian British or Asian Welsh: Bangladeshi
- Asian, Asian British or Asian Welsh: Chinese
- Asian, Asian British or Asian Welsh: Indian
- Asian, Asian British or Asian Welsh: Pakistani
- Asian, Asian British or Asian Welsh: Other Asian
- Black, Black British, Black Welsh, Caribbean or African: African
- Black, Black British, Black Welsh, Caribbean or African: Caribbean
- Black, Black British, Black Welsh, Caribbean or African: Other Black
- Mixed or Multiple ethnic groups: White and Asian
- Mixed or Multiple ethnic groups: White and Black African
- Mixed or Multiple ethnic groups: White and Black Caribbean
- Mixed or Multiple ethnic groups: Other Mixed or Multiple ethnic groups
- Vietnamese
- White: English, Welsh, Scottish, Northern Irish or British
- White: Irish
- White: Gypsy or Irish Traveller
- White: Roma
- White: Other White
- Other ethnic group: Arab
- Other ethnic group: Any other ethnic group

7. What is your work pattern/activity?*

- Paid employment, full time, with at least some work away from home
- Paid employment, part time, with at least some work away from home
- Working from home (full time)
- Working from home (part time)
- Self-employed (business owner, farmer, tradesperson, etc.)
- Attending daycare/school/college/university
- Homemaker (stay at home partner or person with caring responsibilities)
- Unemployed
- Long-term sick-leave or parental leave
- Retired
- Other

8. What is the highest level of formal education/qualification that you have?*

- I have no formal qualification
- GCSE's, O'levels, CSEs or equivalent
- A-levels or equivalent (e.g. Higher, NVQ Level3, BTEC)
- Bachelors Degree (BA, BSc) or equivalent
- Higher Degree or equivalent (e.g. Masters Degree, PGCE, PhD, Medical Doctorate, Advanced Professional Award)
- I am still in education

9. INCLUDING YOU, how many people in each of the following age groups live in your household?*

- 0-4
- 5-12
- 13-18
- 19-44
- 45-64
- 65+

10. How many of the children in your household go to school or day-care?*

- None
- 1
- 2
- 3
- 4
- 5
- More than 5

11. What is your main means of transport?*

- Walking
- Bike
- Motorbike/scooter
- Car
- Public transportation (bus, train, tube, etc)
- Other

12. Did you receive a flu vaccine during the last autumn/winter season? (2024/2025)*

- Yes
- No
- I don't know

13. Are you planning to receive a flu vaccine this autumn/winter season? (2025/2026)*

- Yes, I'm planning to
- Yes, I have got one
- No
- I don't know

14. When were you vaccinated against flu in this season (2025/2026)?*

- Date
- I don’t know/can’t remember

15. What were your reasons for getting a seasonal influenza vaccination this year?*
    Select all options that apply

- I belong to a risk group (e.g, pregnant, over 65, underlying health condition, etc)
- Vaccination decreases my risk of getting influenza
- Vaccination decreases the risk of spreading influenza to others
- It was given to me at my school
- My doctor recommended it
- It was recommended in my workplace/school
- The vaccine was readily available and vaccine administration was convenient
- The vaccine was free (no cost)
- I don't want to miss work/school
- I always get the vaccine
- Other reason(s)

16. What were your reasons for NOT getting a seasonal influenza vaccination in season 2025/2026?*
    *Select all options that apply*

- I am planning to be vaccinated, but haven't been yet
- I haven't been offered the vaccine
- I don't belong to a risk group
- It is better to build your own natural immunity against influenza
- I doubt that the influenza vaccine is effective
- Influenza is a minor illness
- I don't think I am likely to get influenza
- I believe that influenza vaccine can cause influenza
- I am worried that the vaccine is not safe or will cause illness or other adverse events
- I don't like having vaccinations
- The vaccine is not readily available to me
- The vaccine is not free of charge for me
- No particular reason
- Other reason(s)

17. Are you planning to receive a COVID-19 vaccine this season (2025-2026)?

- Yes, I'm planning to
- Yes, I've had one
- No, I’m eligible but not planning to
- No, I’m not in one of the eligible groups
- I don't know

18. Are you planning to receive an RSV vaccine? (pregnant women and those aged 75-79 are eligible)

- Yes, I’m planning to
- Yes, I’ve had one
- No, I’m eligible but not planning to
- No, I’m not in one of the eligible groups
- I don’t know

19. Do you take regular medication for any of the following medical conditions?*
    *Select all options that apply*

- No
- Asthma
- Diabetes
- Chronic lung disorder besides asthma e.g. COPD, emphysema, or other disorders that affect your breathing
- Heart disorder
- Kidney disorder
- An immunocompromising condition (e.g. splenectomy, organ transplant, acquired immune deficiency, cancer treatment)
- I would rather not answer

20. Are you currently pregnant?*

- Yes
- No
- Don't know/would rather not answer

21. Which trimester of the pregnancy are you in?*

- First trimester (week 1-12)
- Second trimester (week 13-28)
- Third trimester (week 29-delivery)
- Don't know/would rather not answer

22. Do you smoke tobacco?*

- No
- Yes, occasionally
- Yes, daily, fewer than 10 times a day
- Yes, daily, 10 or more times a day
- I use e-cigarettes/vapes only
- Dont know/would rather not answer

23. Do you have one of the following allergies that can cause respiratory symptoms?*
    *Select all options that apply*

- Hay fever
- Allergy against house dust mites
- Allergy against domestic animals or pets
- Other allergies that cause respiratory symptoms (e.g. sneezing, runny eyes)
- I do not have an allergy that causes respiratory symptoms

24. Where did you first hear about the FluSurvey?

- Radio or television
- Newspaper or in a magazine
- Internet site (search engine or link)
- Poster
- School or work
- Science events or talks
- Family or friends
- Social media (e.g linkedin, twitter, bluesky)
- UKHSA publications (e.g. reports, blog post)
- Community groups

25. We’re considering including self-swabbing (nose and throat) to test for respiratory viral infections as part of the survey. If this was offered, would you be interested in taking part?*

- Yes, definitely
- Maybe
- No

### FluSurvey weekly questionnaire

* indicates questions shown when symptoms are specified

1. Have you had any of the following symptoms in the past week?

*Please select all options that apply.*

- No symptoms
- Fever
- Chills
- Runny or blocked nose
- Sneezing
- Sore throat
- Cough
- Shortness of breath
- Headache
- Muscle/joint pain
- Chest pain
- Feeling tired or exhausted (malaise)
- Loss of appetite
- Coloured sputum/phlegm
- Watery or bloodshot eyes
- Sticky or itchy eyes
- Nausea
- Vomiting
- Diarrhoea (at least three times a day)
- Stomach ache
- Loss of smell
- Loss of taste
- Nose bleed
- Other, namely:

2. Because of your symptoms, did you use testing at home or were you tested in any health services in relation to these symptoms?

- No, I have not been tested
- Yes, I tested positive for COVID-19
- Yes, I tested positive for Flu
- Yes, I tested positive for RSV
- Yes, I tested positive for more than one of COVID-19, Flu or RSV
- Yes, I tested positive for another infection
- Yes, I was tested but the result was negative
- Don’t know/can’t remember

3. When did the first symptoms appear?*

- Choose date
- I don't know/can't remember

4. When did your symptoms end?*

- Choose date
- I don't know/can't remember
- I am still ill

5. Did your symptoms develop suddenly over a few hours?*

- Yes
- No
- I don't know/can't remember

6. When did your fever begin?*

- Choose date
- I don't know/can't remember

7. Did you take your temperature?*

- Yes
- No
- I don't know

8. What was your highest temperature measured?*

- Below 37.0°C
- 37.0° - 37.4°C
- 37.5° - 37.9°C
- 38.0° - 38.9°C
- 39.0° - 39.9°C
- 40.0°C or more
- I don't know/can't remember

9. Because of your symptoms, did you VISIT (see face to face) any medical services?*

- No
- GP or another health professional at your GP practice
- Hospital accident & emergency department / out of hours service
- Hospital admission
- Other medical services
- No, but I have an appointment scheduled

10. How soon after your symptoms appeared did you first VISIT a medical service?*

- GP or another health professional at your GP practice
- Hospital accident & department/out of hours service
- Hospital admission
- Other medical services

Select the correct number of days:

- Same day
- 1 day
- 2 days
- 3 days
- 4 days
- 5 days
- 6 days
- 7 days
- 8 days
- 9 days
- 10 days
- 11 days
- 12 days
- 13 days
- 14 days
- More than 14 days
- I don't know/can't remember

11. Because of your symptoms, did you contact via TELEPHONE or INTERNET any medical services?*

- No
- GP - spoke to receptionist only
- GP or another health professional at your GP practice
- NHS 111 / NHS 24
- Online NHS Services
- Other
- No, but I have an appointment scheduled

12. How soon after your symptoms appeared did you first contact a medical service via TELEPHONE or INTERNET?*

- GP - spoke to receptionist only
- GP or another health professional at your GP practice
- NHS 111/ NHS 24
- Online NHS Services
- Other Medical Services

Select the correct number of days:

- Same day
- 1 day
- 2 days
- 3 days
- 4 days
- 5 days
- 6 days
- 7 days
- 8 days
- 9 days
- 10 days
- 11 days
- 12 days
- 13 days
- 14 days
- More than 14 days
- I don't know/can't remember

13. Did you take medication for these symptoms?*

Select all options that apply

- No medication
- Pain killers (e.g. paracetamol including Lemsip and Calpol, ibuprofen, aspirin, etc)
- Cough medication (e.g. expectorants)
- Influenza antivirals (e.g. oseltamivir/Tamiflu, zanamivir/Relenza)
- COVID-19 medical treatments such as Paxlovid or sotrovimab
- Antibiotics
- Homeopathy
- Other alternative medicine (essential oil, physiotherapy, etc.)
- Other
- I don't know/can't remember

14. How long after the beginning of your symptoms did you start taking antiviral medication?*

- Same day (within 24 hours)
- 1 day
- 2 days
- 3 days
- 4 days
- 5-7 days
- More than 7 days
- I don't know/can't remember

15. Did you change your daily routine because of your illness?*

- No
- Yes

16. Did you take time off work/school because of your illness?

- No
- Yes
- Not applicable (I am not employed or in education)

17. How long have you been off work/school? (include number of days up to today, if you are still off work/ school) *

- 1 day
- 2 days
- 3 days
- 4 days
- 5 days
- 6 to 10 days
- 11 to 15 days
- More than 15 days

18. Over the course of yesterday, how many people (outside your household) did you approach at a distance lower than 1 metre?

19. On a scale from 0 (bad) to 100 (perfect), how was your health last week?*

*The scale goes from 0 to 100, 100 means the best health status you can imagine and 0 means the worst health status you can imagine.*

*Your answer: Change the value of the slider by dragging the button.*

**The following information is only shown to participants eligible for a self-swab:**

To support our public health surveillance, UKHSA are offering eligible participants with specific symptoms the chance for a free postal respiratory swab test for a range of respiratory viruses. This respiratory swab test involves taking a sample from the back of your throat and from the inside of your nose collected using the same swab. Please note that currently self-swabbing is only open to participants over 18 years old and who live in England.

If you are reporting symptoms on behalf of someone else in your household, please answer these based on their details, and make sure to check that they are willing to participate.

I am interested in more information on this self-swabbing study:

- Yes
- No

If yes:

If you agree, UKHSA will contact you by email with a link to a partner website (Take a Test UK) to provide additional information. Your information will be handled securely by Take a Test UK and limited information will be shared with UKHSA to enable us to identify your sample and test results.

Please note that by agreeing to participate in self-swabbing, you understand that:

- Your test results will be held securely by UKHSA and can be linked back to your survey data
- You will receive certain test results (such as flu and COVID-19) but this test is not intended as a clinical diagnosis or to inform your treatment. If you need urgent results for your individual care, you should not use this service.
- To help us understand risk factors and outcomes of respiratory viruses, your test results will be linked to securely held health data at UKHSA, such as previous immunisations and administrative health records
- These test results will only be sent to you and not to your GP
- You can find full details about how your data will be used, stored, and protected - including the process of withdrawing from the study - in our Privacy Notice

I have reviewed the information provided, including the privacy notice, and agree to take part in this self-swabbing study:

- Yes
- No

Thank you for your filling out the questionnaire. Click submit to send in the responses.

### Feedback survey

1. It was easy to understand the offer of self-swabbing when answering the questionnaire about my symptoms

| 5 point likert scale: | | | | |
| --- | --- | --- | --- | --- |
| Strongly disagree | Disagree | Neutral | Agree | Strongly agree |

2. I understood what testing results would be available if I participated

| 5 point likert scale: | | | | |
| --- | --- | --- | --- | --- |
| Strongly disagree | Disagree | Neutral | Agree | Strongly agree |

3. I remember being provided with information about the testing being offered

| 5 point likert scale: | | | | |
| --- | --- | --- | --- | --- |
| Strongly disagree | Disagree | Neutral | Agree | Strongly agree |

4. I remember reading that my GP would not receive my test results

| 5 point likert scale: | | | | |
| --- | --- | --- | --- | --- |
| Strongly disagree | Disagree | Neutral | Agree | Strongly agree |

5. The process for requesting a kit through Take a Test UK was easy to follow

| 5 point likert scale: | | | | |
| --- | --- | --- | --- | --- |
| Strongly disagree | Disagree | Neutral | Agree | Strongly agree |

6. How long did it take to request a kit through Take a Test UK

Response: Time (minutes)

7. What date did you receive your self-swabbing kit? (leave blank if unknown)
8. I clearly understood the test result being provided

| 5 point likert scale: | | | | |
| --- | --- | --- | --- | --- |
| Strongly disagree | Disagree | Neutral | Agree | Strongly agree |

9. I would consider participating in this self-swabbing for public health surveillance in the future

| 5 point likert scale: | | | | |
| --- | --- | --- | --- | --- |
| Strongly disagree | Disagree | Neutral | Agree | Strongly agree |
